# Counterfactual inference for single-cell gene expression analysis

**DOI:** 10.1101/2021.01.21.21249765

**Authors:** Yongjin Park, Manolis Kellis

## Abstract

Finding a causal gene is a fundamental problem in genomic medicine. We present a causal inference framework that prioritizes disease genes by adjusting confounders without prior knowledge of control variables. We demonstrate that our method substantially improves statistical power in simulations and real-world data analysis of 70k brain cells collected for dissecting Alzheimer’s disease. We identified that 215 causal genes are differentially regulated by the disease in various cell types, including highly relevant genes with a proper cell type context. Genes found in different types enrich distinctive pathways, implicating the importance of cell types in understanding multifaceted disease mechanisms.

## Backgrounds

Single-cell RNA-seq is a scalable approach to measure thousands of gene expression values in hundreds of thousands of cells, sampled from a hundred individuals. As technology becomes mature and economical, single-cell sequencing methods have been used to solve a variety of biological and medical problems, and many large-scale data sets are becoming available to research communities. Unlike previous bulk RNA-seq, single-cell RNA-seq analysis quantifies gene expression changes from a large number of cells, and researchers dare to ask unprecedented questions, which had not been feasible in bulk data analysis. Only a subset of such examples includes cell-level developmental trajectory analysis,^1^ spatial transcriptomics,^2^ regulatory network reconstruction with perturbation,^3^ and variance quantitative trait analysis.^4,5^

Interestingly, some research questions hitherto remain fundamentally attractive since gene expression microarrays^6–8^ and bulk RNA-seq^9–13^ era. Differential expression analysis is such a classical problem. For case-control studies, knowing differentially-expressed genes (DEGs) is often of research and clinical interest. Our primary interest also centres on developing a statistical method for differential expression analysis between different groups of individuals, not between cells. The underlying statistical problem is straightforward. However, finding DEGs from case-control single-cell data poses several challenges in practice. This work seeks to identify and propose an algorithmic approach that resolves two of those challenges from a causal inference perspective.

Firstly, cells are not independently and identically distributed. Instead, cells belong to a particular individual, hierarchically organized, and naturally create “batch” effects (Fig.1a). Cells belonging to the same individual are necessarily affected by the same biological and technical factors. The number of individuals essentially determines the statistical power of DEG discovery in single-cell data. Along the same line, a benchmark comparison demonstrates that existing bulk RNA-seq methods on pseudo-bulk data (using the individual-level aggregate of cells of a particular cell type) still perform decently while correctly controlling false discovery rates.^14,15^ Likewise, for genetic analysis (expression quantitative trait loci), the statistical power of eQTL discovery is primarily determined by the degree of genetic variation across individuals rather than the number of cells per individual.^16^ Nonetheless, differential expression analysis of single-cell RNA-seq is a state-of-the-art and unbiased approach to characterize cell-type-specific transcriptomic changes.

Another challenge stems from the study design of case-control data analysis. In contrast to randomized control trials, most studies are observational, and we have incomplete knowledge of a disease assignment mechanism. Investigators usually cannot make an intervention for practical and ethical reasons. Considering that many complex disease phenotypes occur at the late onset of a lifetime, finding a suitable set of covariates for causal inference is often infeasible as well. Matrix factorization or latent variable modelling can be used to characterize technical covariates or batch effects. However, it is difficult to identify which principal axes of variation capture confounding effects, independently from unknown disease-causing mechanisms. A latent variable model of a single-cell count matrix is frequently used for clustering and cell type annotations, and the resulting latent factors are more suitable for the characterization of intercellular heterogeneity than inter-individual variability.

We present a novel application of a causal inference method as a straightforward approach to improve the statistical power in case-control single-cell analysis while adjusting for unwanted confounding effects existing across heterogeneous individuals. We establish our causal claims in differential expression analysis based on Rubin’s potential outcome framework.^17,18^ Our method is inspired by the seminary work of outcome regression analysis by a matching algorithm.^19,20^ We highlight that our causal inference approach is beneficial in the analysis of disease case-control studies, especially when meta-data for covariates are scarcely available, and covariates may influence both disease status and gene expressions simultaneously. With respect to the underlying causal structural model (disease to gene expression), we seek to identify genes that are differentially-expressed as a result of disease.

## Results

### Problem definition

#### Definition of causal genes

Here, we ask whether a gene is causally affecting or affected by a disease variable but not affected by other technical and biological covariates, which may confound the disease status and gene expressions. In this work, a causal gene is defined as a gene affecting or being affected by a disease status independent of other confounding variables. Although many differentially-expressed genes can be considered a result of disease status for most late-onset disorders, we also acknowledge that aberrant changes on a handful of genes can initiate disease phenotypes. To distinguish causal vs. anti-causal mechanisms, we would need additional perturbation experiments. Alternatively, driver genes can be characterized by mediation analysis using genetic variants as an instrumental variable (Mendelian randomization).^21^

Moreover, concerning cell types and states, we need to assume that cell type fractions are not a mediating factor between the disease and gene expression variables. We found a negligible correlation between cell-type proportions and observed disease status in the study of Alzheimer’s disease.^22^ Under this causal assumption, the stratification procedure for cell types provides a legitimate strategy to control cell-type biases that may impact identifying DEGs. We think there is almost no chance of a “mediation fallacy.^23–25^”

#### Differential analysis on pseudo-bulk expression profiles

We are interested in comparing pseudo-bulk gene expression profiles stratified within each cell type and individual between the case and control samples. Letting *Y*_*gj*_ be a gene expression of a gene *g* on a cell *j* and *S*_*i*_ be a set of cell indexes for an individual *i* ∈ [*n*], we can create a pseudo-bulk expression by aggregating all the expression vectors. We will use *λ*_*gi*_ to generally refer to a pseudo-bulk estimate of a gene *g* on an individual *i*. For instance, we could take an average, *λ*_*gi*_ ≈ ∑_*j*_ *I*{*j* ∈ *S*_*i*_}*Y*_*gj*_/|*S*_*i*_|, or take the total count, 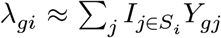. Given the estimate of the *λ* values across *n* individuals, {*λ*_*gi*_ : *i* ∈ [*n*]}, we can construct a hypothesis test that seeks to reject a null hypothesis that the distributions of pseudo-bulk profiles are the same among the case and control individuals (Wilcoxon’s test).

#### Potential outcome framework for single-cell differential expression analysis

In observational data, where the label assignment is not controlled, data matrices of raw {*Y*_*gj*_} and pseudo-bulk count {*λ*_*gi*_} can become confounded with the disease label assignment by unknown biological and technical covariates (Fig. 1b). Such confounding factors obfuscate actual disease-specific effects with other effects of unknown covariates and may lead to false discoveries and dampen the statistical power of differential expression analysis. Rubin’s potential outcome framework^17,18^ seeks to separate the actual disease (or treatment) effects from other effects by asking the following counterfactual questions:

**Figure 1.**
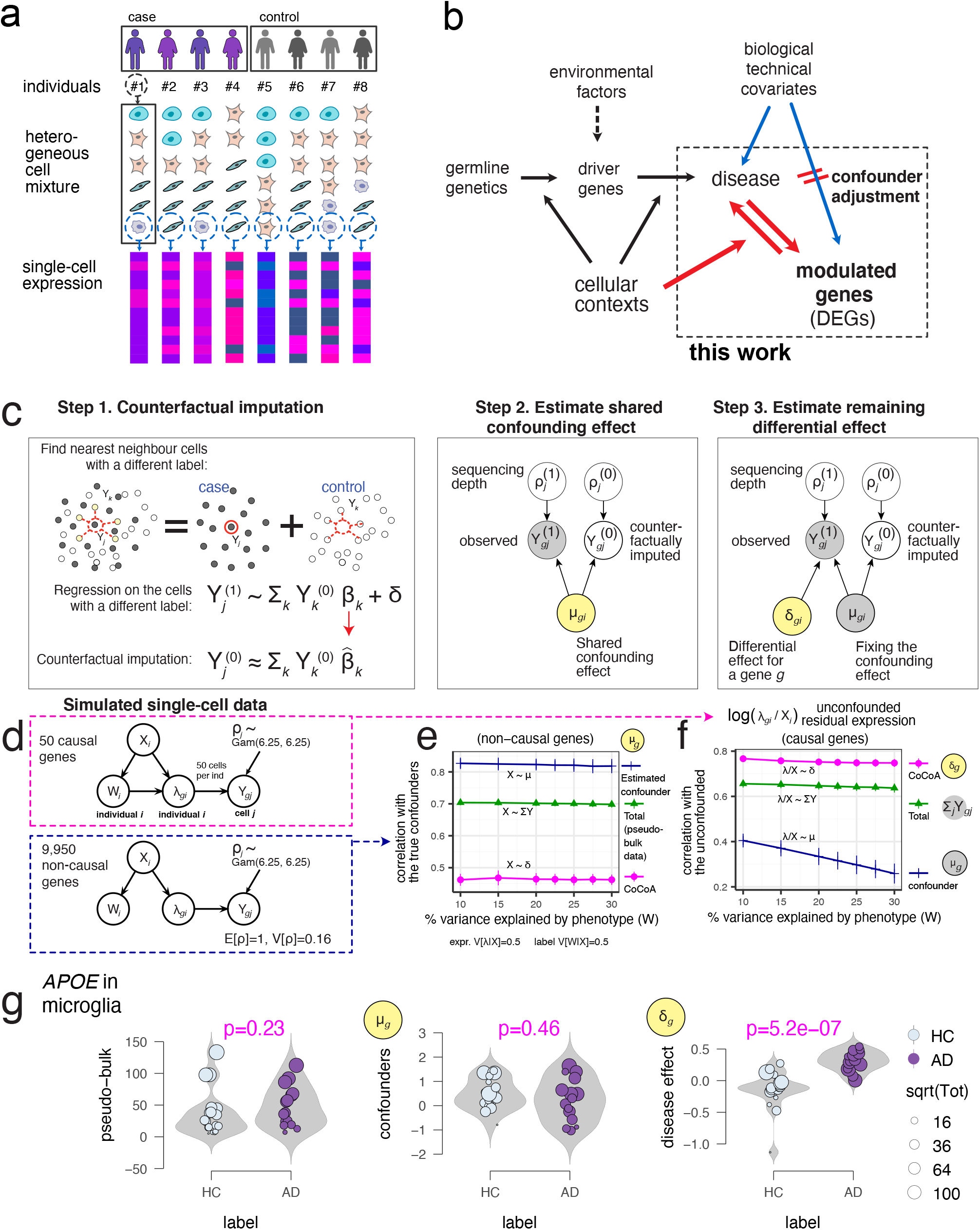
Counterfactual confounder adjustment (CoCoA) for single-cell differential gene expression analysis. **(a)** Hierarchical (nested) structure of single-cell gene expression data. We have tens of individuals for one case-control study. Each individual (*i*) contains a heterogeneous mixture of multiple cell types. Single-cell technology measure a thousand genes on each cell (*j*). **(b)** This work seeks to address a specific causal inference problem of genomics research. We seeks to prioritize genes causally-modulated by a disease status, not the genes affecting the predisposition and risk of disease development. **(c)** Overview of CoCoA approach (see Methods for details). *Y* : gene expression matrix. *Y* ^(0)^: counterfactual data with disease *W* = 0, *Y* ^(1)^: counterfactual data with disease *W* = 1. *β*: Poisson regression coefficient. *δ*: residual effect. *ρ*: sequencing depth. *μ*: shared confounding effect. **(d)** Data generation scheme for simulation studies. We simulate 50 causal and 9,950 non-causal genes with or without disease-causing mechanisms (an edge between *W* and *λ*). *W*_*i*_: disease label assignment for an individual *i. X*_*i*_: confounding effects for an individual *i. λ*_*gi*_: unobserved gene expression for a gene *g* of an individual *i* as a function of *X* and *W*. *Y*_*gj*_: realization of cell-level gene expression of a gene *g* with a cell *j*-specific sequencing depth *ρ*_*j*_ (stochastically sampled from Gamma distribution). Here, we simulated five different *X* variables. CoCoA accurately estimates shared confounder variables (*μ*_*g*_), showing a significantly-higher level of correlation with true confounding effects on non-causal genes than a pseudo-bulk analysis. CoCoA accurately estimates disease-causing effects (*δ*_*g*_), showing high correlation with true differential effects on causal genes. **(g)** Illustration of CoCoA approach on the *APOE* in microglia example. HC: Health Control. AD: Alzheimer’s disease. *μ*: shared confounding effect; *δ*: residual differential effect. For a clear visualization, we omitted samples (individuals) with zero read count observed on *APOE* gene in the microglial cell type.

- What would be a gene expression if an individual had not been exposed to a disease?
- What would be a gene expression if an individual had a disease?

In our pseudo-bulk analysis context, we are interested in estimating the following quantities:

- 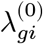: What would be the pseudo-bulk expression of a gene *g* if an individual *i* had not been exposed to a disease?
- 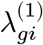: What would be the pseudo-bulk expression of a gene *g* if an individual *i* had exposed to a disease?

In a binary case-control study, we observe one of the values for each individual while the other side is left unobserved (denoted by the “?” mark). Letting *W*_*i*_ ∈ {0, 1} be a disease label assignment variable for an individual *i*, only a part of potential gene expressions are made directly observable from data:

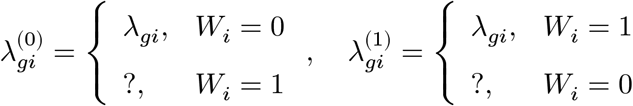

At the cell level (*∀j* ∈ *S*_*i*_), we have the same structure:

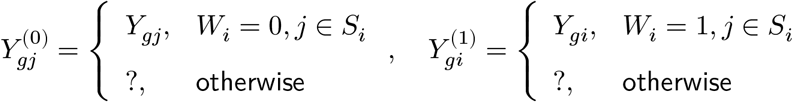

If both sides of the potential expression, 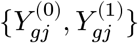, were known, we would be able to estimate the disease effect on a gene *g* for each individual by comparing pseudo-bulk profiles 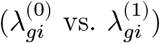 constructed from the potential single-cell gene expressions. The ultimate goal of causal inference in Rubin’s potential outcome framework is to impute the missing part of potential outcomes since a comparison between the case and control becomes straightforward on the imputed data.

#### The definition of a confounding variable and causal assumptions

We define that a variable can confound a disease label (*W*) and gene expressions (*λ* and *Y*) if (1) it is associated with the disease and gene expression variables; (2) it is still associated with the expression even after conditioning on the disease label.^26^ Unless we adjust/stratify a sufficient set of confounding variables, gene expression changes observed between the case and control samples are not necessarily the causal effect of disease mechanisms.

It is crucial to state casual assumptions to proceed with our causal inference:

- *Stable individual disease effect*: We assume that the potential expressions of an individual *i*, namely 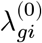 and 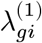, are not affected by the expressions of other individuals {*i*′ ∈ [*n*] : *i*′ ≠ *i*}.
- *Conditional independence of the potential expression and disease exposure*: For a non-causal gene, by definition, gene expressions are independent of disease status. Therefore we do not need an assumption on this matter. However, for a causal gene, we assume that potential (counterfactual) gene expressions are independent of a disease label conditioning on a sufficient set of confounding variables. In other words, genes differentially regulated for a diseased individual would not have been aberrantly expressed if this individual had not developed the disease.
- *Overlap of confounding effects between the case and control*: Within a stratum of individuals, homogeneous with respect to confounding variables, we have both the case and control subjects with non-zero probability. In the single-cell analysis, we assume disease and non-disease cells simultaneously exist in a homogeneous group of cells stratified by confounding factors.

#### CoCoA for single-cell differential expression analysis

The purpose of our counterfactual confounder adjustment (CoCoA) for single-cell gene expression analysis (Fig. 1c) is to impute the missing part of potential outcomes of single-cell profiles (step 1), propagate the imputed results to the pseudo-bulk estimation, and decompose the total pseudo-bulk profiles into the confounding (step 2) and differential effects (step 3). Using a single-cell gene expression matrix, {*Y*_*gj*_ : *g* ∈ genes, *j* ∈ cells}, we want to estimate two types of pseudo-bulk data: (1) the estimated confounders, {*μ*_*gi*_ : *g* ∈ genes, *i* ∈ individuals}, and the residual differential effects, {*δ*_*gi*_ : *g* ∈ genes, *i* ∈ individuals}. In other words, we want to estimate the decomposition of pseudo-bulk data, such as *λ*_*gi*_ = *μ*_*gi*_*δ*_*gi*_. Briefly, the algorithm proceeds as follows: (1) We seek to estimate (or impute) counterfactual measurement of single cells’ expression by matching cells in a particular condition with neighbouring cells in the opposite conditions. The distance between cells was calculated on the top principal component axes. (2) Having paired sets of observed and counterfactual single-cell data, we estimate the mean expression of genes shared across two opposite conditions in Bayesian posterior inference. We treat them as putative confounding factors. (3) While holding the estimated confounding effects fixed, we measure the conditional (or residual) mean effect on the observed cells. We refer the readers to Materials and Methods for technical details.

To demonstrate how CoCoA actually works, we simulated a single-cell data matrix consisting of 10,000 genes and 40 individuals (Fig. 1d). Each individual contains 50 cells on average; 50 of the 10,000 genes are causally affected by disease labels (*W* → *λ*) and confounding factors (*X* → *λ*). The other genes are only affected by confounding factors (*X* → *λ*); we introduced five confounding variables {*X*_*ik*_ : *i* ∈ individuals, *k* ∈ [5]}, and a linear combination of these *X* variables introduces biases on *W*_*i*_ and *λ*_*gi*_. Here, we set the variance of *λ* explained by confounding variables *X* to 0.5 and the variance of disease label *W* explained by the same confounding variables to 0.5, but we varied the true disease variability between 0.1 to 0.3 on 50 causal genes (*W* → *λ*; the x-axes of Fig. 1e-f). As expected, on non-causal genes (Fig. 1e), we found a strong correlation between the estimated confounding effects (*μ*_*gi*_) and true confounding effects (the linear combination of *X*_*ik*_ variables), which is far greater than the correlation with the estimated differential effects (*δ*_*gi*_). We also observed that the unconfounded pseudo-bulk data (removing the effects of *X* variables) are correlated with the estimated differential effects (*δ*_*gi*_), consistently not affected by the change of disease variability (Fig. 1f).

As an example, we demonstrate the effectiveness of our approach in the case of *APOE* gene measured in microglia samples (Fig. 1g). For better visualization, we removed an individual with only a single read was observed on the *APOE* gene. In pseudo-bulk data analysis with 39 individuals, it appears that the total expression values are only mildly up-regulated in disease subjects (Wilcoxon p = 0.23) even though *APOE* over-expression is one of the most frequently observed hallmark of Alzheimer’s disease (AD). We also found that the shared confounding factors across the case and control exhibit almost no apparent correlation with the disease label (p=0.46). After adjusting the confounders on the data, we recover a significant correlation of *APOE* gene expressions with AD status (p=5.2 ×10^−7^).

### Simulation experiments

#### The design of simulation experiments

We evaluated the performance of CoCoA in downstream differential expression analysis using a simulated single-cell data matrix (*Y*) of 10,000 genes with 40 individuals with 50 causal and 9,950 non-causal genes (Fig. 2a). We introduced two types of total five covariates on the individual-level expressions *λ*_*gi*_, explicitly designating confounding variables *X* and non-confounding batch effect *B*. By definition, confounding factors *X*_*ik*_ affect the label assignment *W*_*i*_ and the individual-level mean values *λ*_*gi*_. For a causal gene *g, λ*_*gi*_ values are determined by the disease label *W*_*i*_, the confounders *X*_*ik*_, and the batch effect variables *B*_*il*_; for a non-causal gene *g*, there is no contribution from the disease variable. In each simulation experiment, we specify the following parameters:

**Figure 2.**
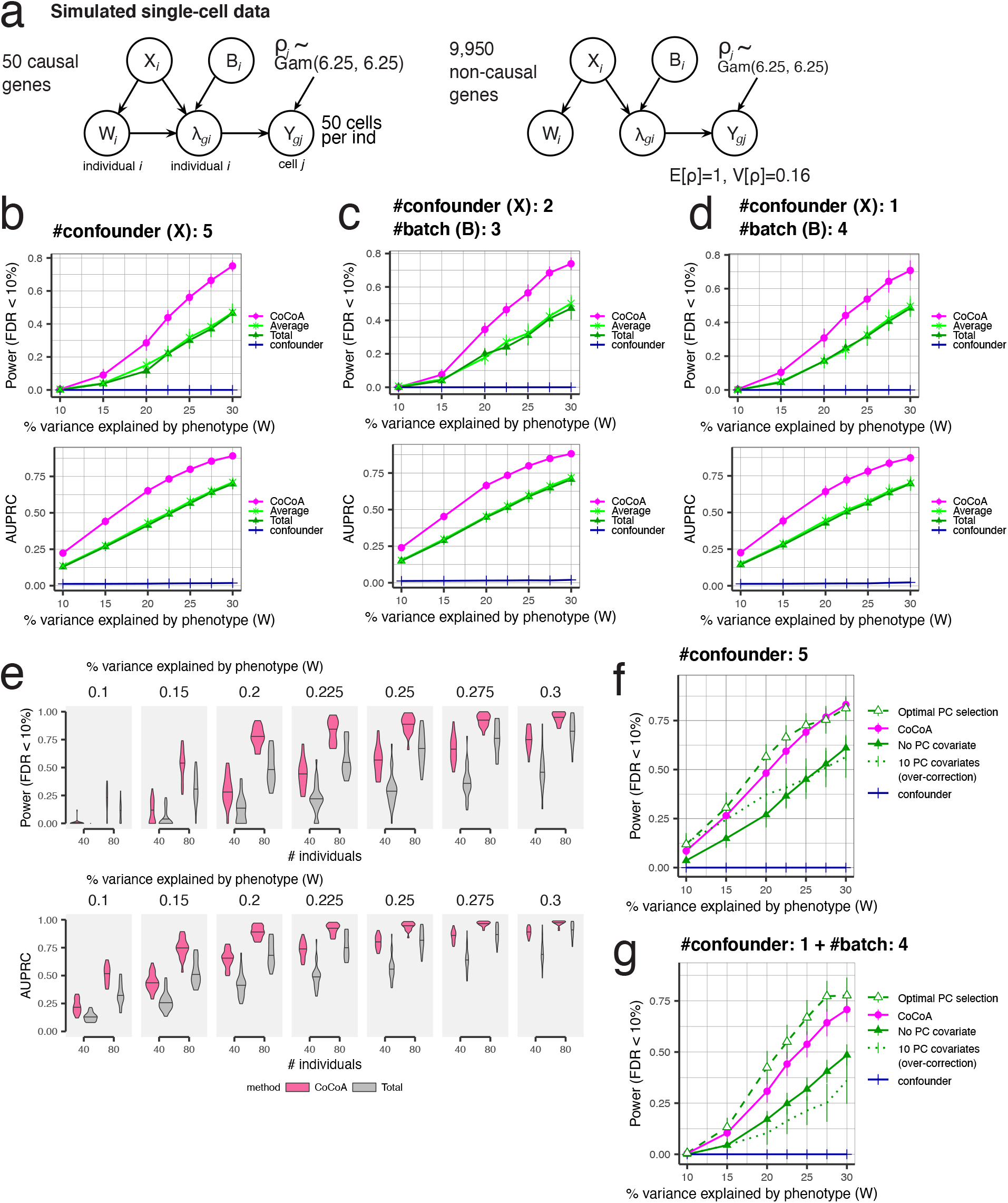
Simulation experiments. Extensive simulation experiments confirm that CoCoA effectively adjusts existing confounding effects and improves statistical power of differential expression analysis. **(a)** Data generation scheme for simulation experiments. We simulate 50 causal and 9,950 non-causal genes with or without disease-causing mechanisms (an edge between *W* and *λ*). *W*_*i*_: disease label assignment for an individual *i. X*_*i*_: confounding effects for an individual *i. λ*_*gi*_: unobserved gene expression for a gene *g* of an individual *i* as a function of *X* and *W*. *Y*_*gj*_: realization of cell-level gene expression of a gene *g* with a cell *j*-specific sequencing depth *ρ*_*j*_ (stochastically sampled from Gamma distribution). Here, we simulated total five covariates consisting of confounding (*X*) and batch effect variables (*B*). **(b)** Simulation results when all the five covariates are confounding disease label assignment and gene expression values. *CoCoA*: Wilcoxon’s ranksum test using individual-specific confounder-adjusted gene expression values *δ*_*gi*_ (the step 3 of Fig. 1c); *total*: pseudo-bulk expression aggregated within each individual; *average*: pseudo-bulk expression averaged over cells within each individual; *confoudner*: the estimated confounding effect *μ*_*gi*_ (the step 2 of Fig. 1c). *AUPRC* : area under precision recall curve (numerically integrated by `DescTool` implemented in R).28 **c)** Simulation results when only one covariate confounds disease label assignment and gene expression values, and the other four variables are affecting gene expression as batch effects. **(d)** Can sample size help? Simulation results compare the statistical power of differential expression analysis conducted on two different sizes of simulation studies (N=40 vs. N=80). **(e)** CoCoA avoids over-correction of expression variation and attains nearly optimal performance in differential analysis when there is only one confounding factor and the other four factors correspond to batch effects.

- 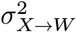: the variance of *W* explained by the covariate *X*.
- 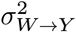: the variance of log *λ* explained by the disease assignment *W*.
- 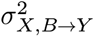: the variance of log *λ* explained by the covariates *X* and *B*.
- *d*_*C*_: the number of confounding variables (from 1 to 5).
- *d*_*B*_: The number of non-confounding batch effects (from 0 to 4).

For each individual *i* ∈ [*n*], we first sample covariates *X*_*ik*_ ∼ *𝒩*(0, 1) for *k* ∈ [*d*_*C*_] and *B*_*il*_ ∼ *𝒩*(0, 1) for *l* ∈ [*d*_*B*_]. Given the *X* matrix, we sample the parameter vector *α* required to introduce biases on *W* and the residual error vector *ϵ*_*W*_ from isotropic Gaussian distributions and adjusted the scale of the error vector to have the simulation proportion of variance matched with the prescribed *σ*^2^ value, i.e., 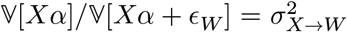. We generate a binary label assignment for an individual *i* by flipping a coin:

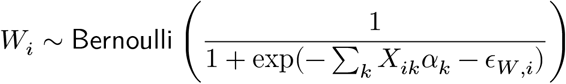

Combining these values, we construct the mean values of a gene expression *g* for an individual *i* by a generalized linear model:

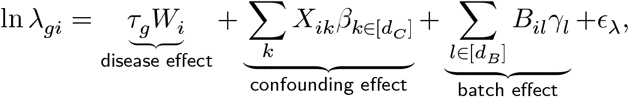

where the covariate effect 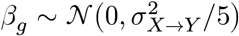, gene-level causal effect 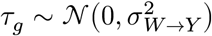, and the residuals 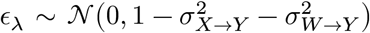. Using the individual-level mean values *λ*, we stochastically generated 50 cells per individual, multiplying the individual-level average expressions with random sequencing depth (*ρ*), sampled from *ρ* ∼ Gamma(6.25, 6.25). For each cell *j*,

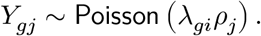

#### Counterfactual adjustment of pseudo-bulk data improves statistical power

We repeated our experiments 30 times for all the different configurations and summarized the performance by the area under the precision-recall curve (AUPRC) and the statistical power when the false discovery rate was controlled at 10% (Fig. 2b-d). Once we have estimated different types of pseudo-bulk data, we ranked genes based on Wilcoxon’s rank-sum test^27^ implemented in R and constructed receiver-operating and precision-recall curves to calculate the power and AUPRC using `DescTool`^28^ implemented in `R.` Extensive simulation results can be found in the supplementary website (https://ypark.github.io/cocoa_paper/result_simulation.html) along with the source code repository (https://github.com/ypark/cocoa_paper).

Here, we show and compare the performance of differential analysis conducted on the four different pseudo-bulk data:

- CoCoA: individual-level disease effects *δ*_*gi*_ estimated by CoCoA algorithm (Fig. 1c, step 3).
- Confounder (estimated): the remaining confounder effects *μ*_*gi*_, also esimated by CoCoA algorithm (Fig. 1c, step 2)
- Average: arithmetic mean of cell-level expressions within each individual 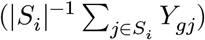.
- Total: summation of cell-level expressions within each individual 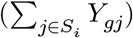.

We found that CoCoA effectively adjusts the simulated confounding effects and markedly improves statistical power and AUPRC of differential expression analysis. We also confirmed that the confounder effects that we corrected show no association with disease labels. Unlike different types of covariates (confounding and batch effects) create confusion to an algorithm in selecting factors to be adjusted in a matrix factorization-based analysis, CoCoA performed robustly, showing only a modest reduction of power on large batch effects.

We further investigated whether the bias introduced by confounding effects can be overcome by doubling the number of individuals (Fig. 2e). Because of imperfect confoundedness (50% of variability), we found that larger sample size can improve the accuracy of differential analysis. However, the performance gap between the CoCoA and pseudo-bulk analysis by total expression sustains in almost all cases.

Lastly, we considered a hypothetical scenario where principal components of a gene expression matrix are optimally selected to characterize confounding factors (Fig. 2f-g). CoCoA closely attains the optimal level of statistical power when all the covariates act as a confounding factor (Fig. 2f). However, if there exist other non-confounding factors, CoCoA alone may not reduce all the other types of unwanted variation in a single-cell data matrix (Fig. 2g).

#### Case study: Finding cell-type-specific causal genes in Alzheimer’s disease

We reanalyzed published single-nuclei RNA-seq (snRNA-seq) data of 48 individuals in postmortem brain samples.^22^ To our knowledge, this is one of the largest snRNA-seq data on case-control disease studies. Of the 48 individuals, we included 40 individuals for differential expression analysis because we found no case-control disease labels on the reaming eight individuals.

#### Cell type annotations of 70,634 cells

We directly annotated the cell types of these 70,634 cells using the list of cell-type-specific genes provided by the PsychENCODE project.^29^ Of the total 2,648 PsychENCODE marker genes, we used 1,726 genes expressed in this data set as features (Supplementary Fig. 1). We identified the eight cell-type clusters of cells while estimating a mixture of von Mises-Fisher distributions, measuring cells’ likelihood to centroids by angular distance (see Methods). We found that this gene-to-cell-type membership information was sufficient enough to distinguish eight cell types. These eight cell types include expiatory (Ex) and inhibitory neurons (In), oligodendrocytes (Oligo), oligodendrocyte progenitor cells (OPC), microglia, astrocytes (Astro), pericytes (Per), and endothelial cells (Endo). We found that our annotation almost perfectly agrees with the original paper’s cell type annotation (Supplementary Fig. 2). We also found that cell types correspond to unique cell clusters after BBKNN (batch-balancing k-Nearest Neighbour)^30^ preprocessing (Fig. 3a), showing no apparent bias induced by other demographic and pathological variables (Supplementary Fig. 3). Moreover, we further dissected four different cortical layer-specific cell types for excitatory neurons (Fig. 3b) and four different subtypes for inhibitory neurons (Fig. 3c) using a refined set of marker genes provided by previous single-nucleus analysis.^31^

**Figure 3.**
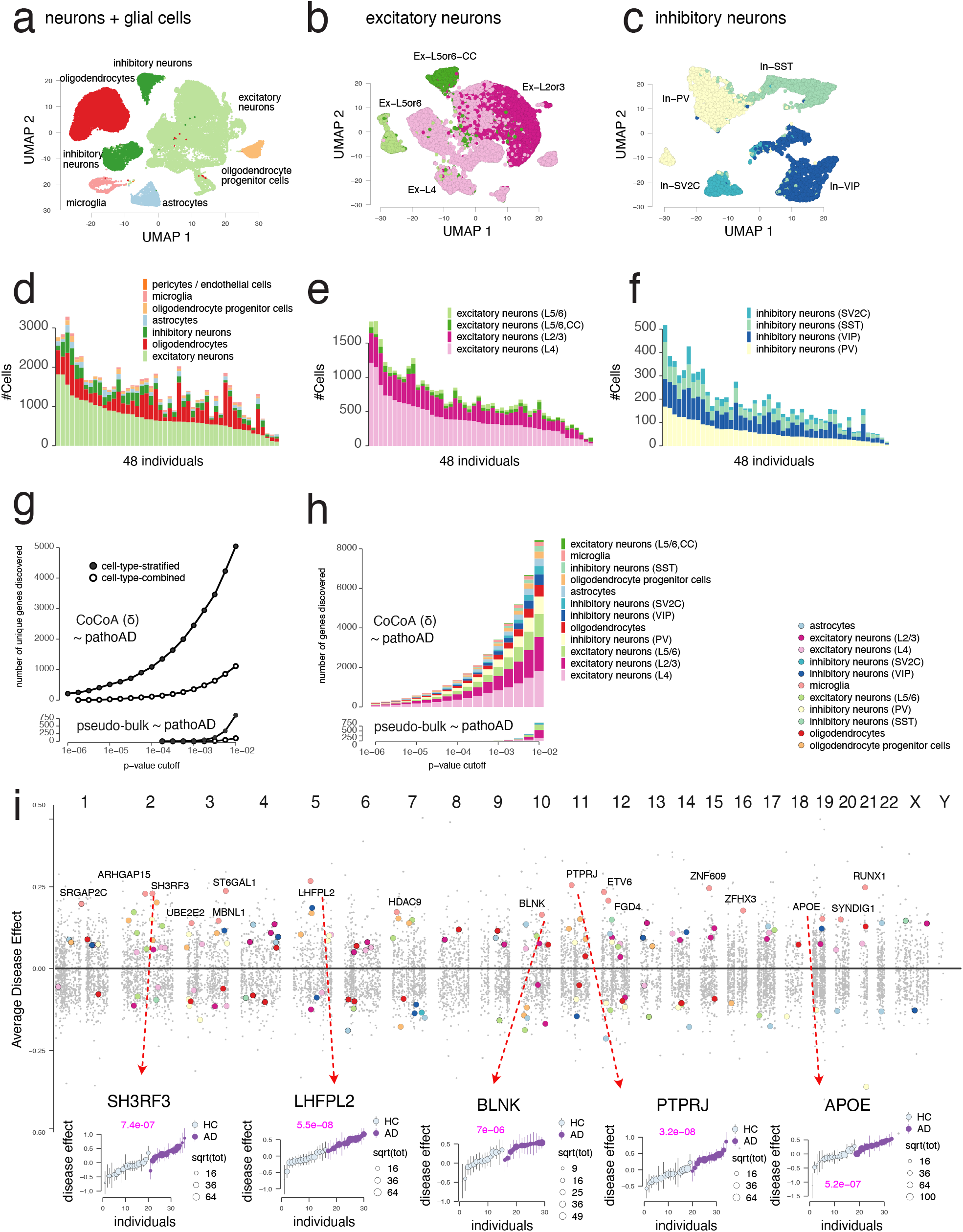
The case study of Alzheimer’s disease snRNA-seq profiles.^22^. Cell-type-stratified approach improves statistical power and interpretation of differential expression analysis. **(a)** UMAP projection of the major cell types. **(b)** UMAP projection of the excitatory neuron subtypes. **(c)** UMAP projection of the inhibitory neuron subtypes. **(d)** Cell type decomposition of the major cell types across 48 individuals. **(e)** Cell type decomposition of the excitatory neurons across 48 individuals. **(f)** Cell type decomposition of the inhibitory neurons across 48 individuals. **(g-h)** Cell-type-stratified CoCoA approach improves statistical power and identifies genes in a wide spectrum of the cell types. **(i)** A genomic view of genes strongly-modulated by AD pathology (coloured). *Top panel*: genomic location of cell-type-specific causal genes; *bottom panels*: five representative examples of the most significant genes.

We notice a wide spectrum of cell-type variability across 48 individuals, both in terms of the number of cells and proportions (Fig. 3d). The Mathys *et al*. data contain on average 1,444 cells per individual, of which 50.65% cells stem from Ex (N=726 ± 382 SD), 12.71% cells from In (N=182 ± 107), 25.72% cells for Oligo (N=380 ± 252), 3.56% cells from OPC (N= 54 ± 34), 2.43% cells from Microglia (N= 33 ± 24), 4.94% cells from Astro (N= 69 ± 46), 0.1% cells from Endo and Per. We have 726 excitatory neurons per individual (Fig. 3e), of which 52.62% (N=399 ± 250) cells from the layer 4, 33.1% (N=233 ± 108) cells from the layer L2/3, 8.26% (N= 50 ± 28) cells from the cortical layer L5/6 (CC), 6.03% (N= 44 ± 31) cells from the layer 5/6. We have 182 inhibitory neurons per individual (Fig. 3c), consisting of 32.9% (N=59 ± 34) cells from inhibitory neurons (VIP), 31.72% (N=59 ± 40) cells from inhibitory neurons (PV), 24.51% (N=43 ± 26) cells from inhibitory neurons (SST), 11.1% (N=22 ± 17) cells from inhibitory neurons (SV2C).

#### Cell type stratification improves the statistical power and interpretation of differential expression analysis

We investigated the impact of such a high level of cell-type heterogeneity on subsequent differential expression analysis. Tissue-level bulk RNA-seq analysis data can be arguably thought of as an aggregate of single-cell-level expressions. If genes were similarly affected by the disease phenotype in most cell types, we would expect the bulk-level associations to be similar, and cell-type-stratified analysis would benefit less than more of stochasticity–fewer cells per individual. On the other hand, if most disease-responsive genes act through a cell-type-specific mechanism, cell-type-stratified data analysis will improve statistical power and render better biological interpretations in genomics analysis.

Using these cell type annotations, we constructed cell-type-stratified pseudo-bulk data for all the genes and individuals in each cell type independently, treated them as a gene expression matrix, and tested associations of genes with AD status. We also constructed the pseudo-bulk profiles by combining all the cells in each individual, ignoring the cell type annotations, and carried out the same association analysis. It is clearly shown that the number of discoveries (unique genes) dramatically increase with cell-type-specific stratification steps in both studies using CoCoA and total expression profiles (Fig. 3g). Considering the variety of cell types in each p-value cutoff, such cell-type-specific mechanisms are likely to remain hidden in bulk, combined differential analysis but better revealed after taking into account cell type heterogeneity (Fig. 3h).

### Disease status modulates the cell-type-specific gene expressions

#### 215 genes are differentially-regulated with AD pathology

We prioritized genes based on testing a hypothesis that the pseudo-bulk profiles processed by CoCoA are differentially ranked by AD pathology (Wilcoxon’s ranksum test).^27^ We conservatively adjusted putative confounding effects with (100-nearest neighbour search) in a spectral space constructed by 50 principal components. Controlling the false discovery rate (FDR^32^) < 1%, we found 1,648 genes (11.68% of 14,106), consisting of 672 genes found in Ex-L4, 522 in Ex-L2/3, 297 in Ex-L5/6, 210 in In-PV, 98 in Oligo, 84 in Microglia, 80 in Astro, 57 in In-VIP, 49 in OPC, 11 in In-SST, and 4 in In-SV2C. Controlling family-wise error rate (FWER^33^) at 1%, we found a total of 215 genes (1.52%), which consist of 55 genes found in Ex-L4, 39 in Ex-L2/3, 28 in Ex-L5/6, 28 in Oligo 24 in In-PV, 19 in Microglia, 19 in Astro, 9 in OPC, 7 in In-VIP, and 3 in In-SST.

We confirmed that the CoCoA procedure did not introduce a systematic bias by shrinking variance on the case or control samples (Supplementary Fig. 6). We tested our method on four different phenotypes using twelve cell-type-specifically confounder-adjusted profiles and cell-type-sorted pseudo-bulk data. Moreover, visual inspection of the p-value distributions for Wilcoxon’s tests suggests no apparent inflation/deflation in our multiple hypothesis testing (Supplementary Fig. 7).

In addition to the non-parametric ranksum test, we propose a model-based Wald statistic for an individual-level test (for each gene *g* and an individual *i*), namely *Z*_*gi*_ = 𝔼[ln *δ*_*gi*_]/𝕍[ln *δ*_*gi*_], and the group-wise average disease effect size (ADE) and standard error (SE) for each gene *g*:

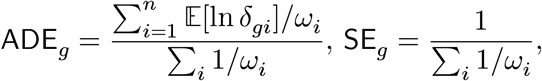

where *ω*_*i*_ = 1/𝕍[ln *δ*_*gi*_] for brevity (the method). We found that gene-level ADE values are marginally independent of average confounding effects (the top panels of Supplementary Fig. 8). However, we confirmed that average disease effects on the disease samples (ADD) generally align well with the average disease effects computed on the control samples (the bottom, Supplementary Fig. 8).

The false sign rate (FSR) of these Bayesian estimates of ADE and SE can be controlled by an empirical Bayes procedure, such as `ashr`.^34^ Controlling the FSR of ADEs and FDR of the ranksum tests both below 1%, we found a total of 1,330 AD genes (1,669 gene and cell type pairs) and an average of 152 (± 206) genes per cell type. Of them, we highlighted 182 genes sampled at most 20 genes within each cell type (Fig. 3i) and annotated 17 genes specifically acted in the microglia. We found multiple lines of independent evidence to corroborate the causal role of these genes.

Of these top AD genes found in microglial cells, we highlight five genes, including *APOE*, sh wing gene expressions up-regulated clearly among the AD individuals (the bottom panels of Fig. 3i). *SH3RF3* gene has been found significantly associated with the age at onset of AD in the family-based genome-wide association studies.^35^ Interestingly, regarding Parkinson’s disease, another neurodegenerative disorder, genetic variants located in *LHFPL2* have been associated with accelerated onset of the disease by 8 to 12 years.^36^ *BLNK* plays a key regulatory role in well-known microglia-specific *TREM2* signaling pathway^37^ and has been proved to be up-regulated with the increase of amyloid *β* protein.^38^ To some degree, conditional genetic analysis suggested that *PTPRJ* is a link to explain pleiotropy between late-onset AD and major depressive disorder.^39^

The full statistics and the visualization of representative examples are shared in the supplementary website (https://ypark.github.io/cocoa_paper/result_AD.html).

#### Gene ontology analysis characterizes a variety of cell-type-specific pathways in AD

We sought to characterize cell-type-specific mechanisms potentially perturbed by 1,077 (± 1,122) significant AD genes found in each cell type (FDR < 20%) using goseq^40^ package. Gene ontology (GO) enrichment analysis shows that DEGs identified in different cell types indeed influence markedly different biological mechanisms. By visual inspection, we can identify cell-type-specific modules of the enriched GO terms in the biological process category (Fig. 4b-d). For instance, up-regulated AD genes found in excitatory neurons are highly enriched in neurodevelopmental pathways, such as “modulation of chemical synaptic transmission,” “regulation of trans-synaptic signalling.” However, microglial DEGs are mostly found in immune-related activities, such as “interferon-gamma-mediated signalling pathway” and “regulation of lymphocyte-mediated immunity,” and oligodendrocyte DEGs enrich terms reflect the functional role of the cell type such as “myelination” and “axon ensheathment.” For the GO terms in the cellular component category, DEGs found in neurons over-represent synapse and axon, but glial cell-type-specific DEGs highlight cell-cell junction and focal adhesion (Fig. 4e). DEGs found in neurons generally participate in ion channel activities, but we noticed that microglial DEGs are highly relevant to Rho GTPase and cadherin binding activities (Fig. 4f).

**Figure 4.**
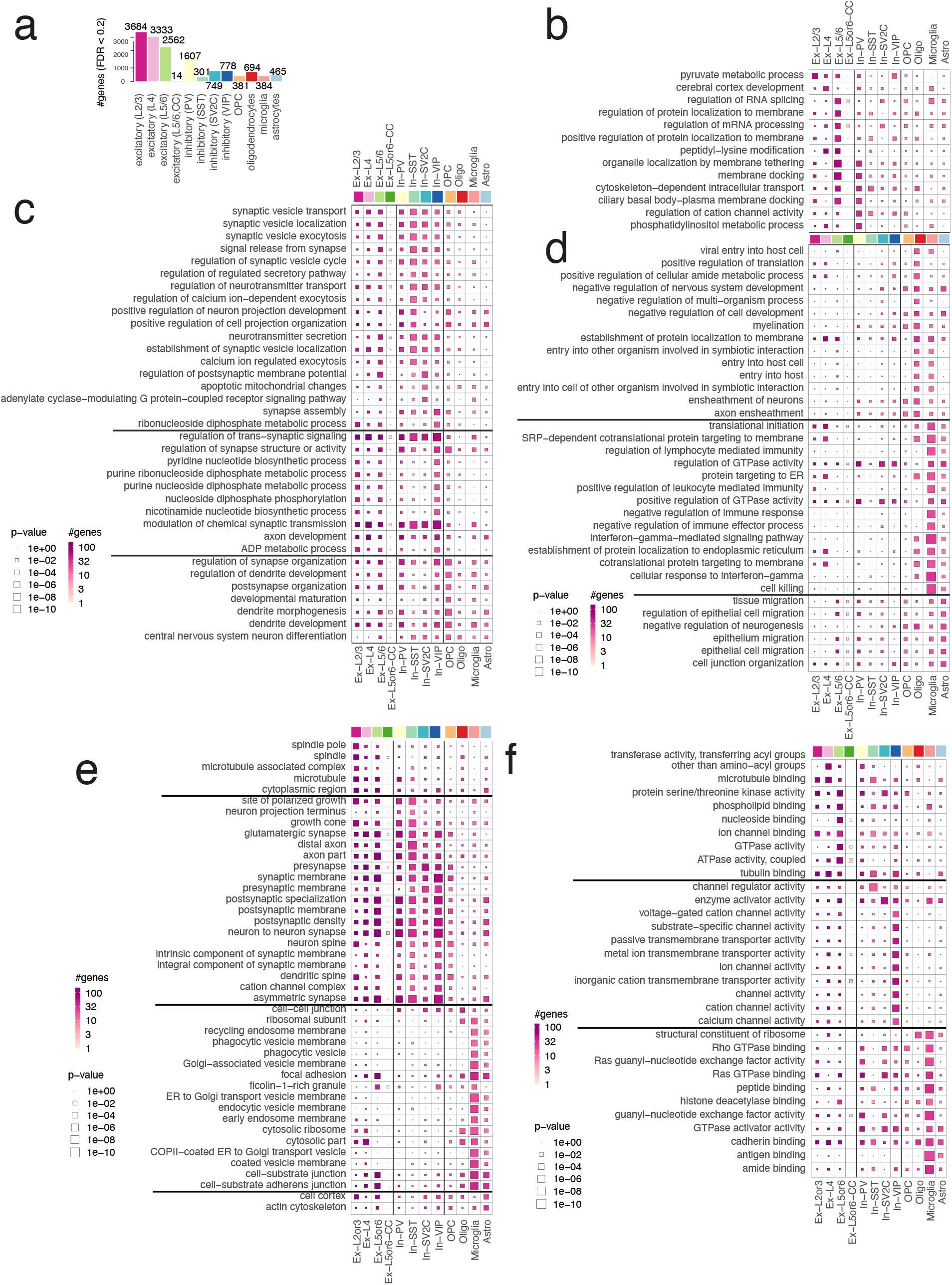
Genes modulated by AD pathology highlight disease mechanisms in relevant cell types in gene ontology enrichment analysis. *Ex*: expiatory neurons; *In*: inhibitory neurons; *Oligo*: oligodendrocytes; *OPC* : oligodendrocyte progenitor cells; *Mic*: microglia; *Astro*: astrocytes. Each box is scaled proportional to the level of enrichment significance (p-value), and the colour gradient marks the number genes overlapped in each keyword and cell type. **(a)** The number of significant genes modulated by AD pathology. **(b-d)** Top gene ontology terms in biological process over-represented by cell-type-specific AD genes. **(e)** Top gene ontology terms in cellular component over-represented by cell-type-specific AD genes. **(f)** Top gene ontology terms in molecular function over-represented by cell-type-specific AD genes.

The full statistics of gene ontology enrichment analysis are available through the supplementary website (https://ypark.github.io/cocoa_paper/result_AD_GO.html).

## Discussion

We addressed a subset of a causal inference problem that emerges in disease studies. We sought to characterize and estimate the average causal effect of genes between the case and control individuals from observational single-cell data. Delineating confounding and non-causal factors from causal effects is a crucial step to many genomics problems. Not to be trapped in circular reasoning (identifiability issue), the genomics community has been using so-called control genes and samples to extract factors shared in both control and discovery data^41–43^. One of the steps in our algorithm enjoys a similar idea, but there is no need to prescribe control cells or genes for our purposes. Along the same line, only if control features were known a priori, contrastive principal component analysis^44^ could pick out non-causal factors in its latent space. Likewise, only if nuisance variables are independently observed, the variational fair autoencoder model^45^ project cells onto unconfounded (“fair”) latent space.

Our method builds on the outcome regression facilitate by a matching algorithm.^19,20^ Like most existing single-cell analysis pipelines, finding reliable k-nearest neighbour cells is a crucial step. If some cells in one condition were poorly matched with other cells in the opposite condition, failing to capture a shared component of confounding effects, our analysis might not work as expected. However, we want to emphasize that a failure of the matching step does not lead to an over-correction of pseudo-bulk data. It is important to understand and reliably quantify to what degree a cell-cell matching procedure can address the intrinsic and another technical variability of a single-cell RNA-seq data matrix.

A sparsity of single-cell data still casts a wide range of modelling questions. As we only consider the average effect within each individual, and we take a simple model that is just enough to capture our estimands. We ignored the notion of zero-inflation since we treat single-cell data as a count matrix, not being transformed by logarithm^46^. However, future research can take advantage of more sophisticated modelling of the individual- and cell-level observations^47^, perhaps involving latent variables for representational learning.

## Conclusions

We present a causal inference method that identifies and removes putative confounding effects from single-cell RNA-seq data so that the subsequent differential expression analysis can become unbiased and gain more statistical power. We have empirically shown that CoCoA improved the downstream data analysis in extensive simulation experiments. We also demonstrated in real-world snRNA-seq data that the CoCoA approach was necessary to reveal both well-established and novel causal genes in AD. Our work is the first application of counterfactual inference to single-cell genomics to the best of our knowledge. We expect that many existing inference methods and models can be reformulated in the same causal inference framework. More broadly, we believe that causal inference methods can improve the interpretation of genomics analysis and ultimately benefit translation researches.

## Methods

### Preliminary modelling of single-cell RNA-seq counting data

#### Individual-level gene expression quantification

We describe the single-cell RNA-seq data-generating process in a Poisson-Gamma hierarchical model. For each individual, we measure thousands of gene expression on nearly a thousand cells. Here, we denote each individual, gene, and cell by index *i, g, j*, respectively. We model the expression count *Y*_*gj*_ of a gene *g* in a cell *j* follows Poisson distribution with the composite rate parameter, *λ*_*gi*_*ρ*_*j*_, where *λ*_*gi*_ quantifies the gene’s mean activity in the corresponding individual *i*, and *ρ*_*j*_ accounts for the sequencing depth of a cell *j*. More precisely, we define the likelihood of *Y*_*gj*_:

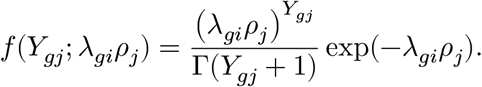

We assume the gene and cell parameters, *λ, ρ*, follow a conjugate prior distribution (Gamma); more precisely, we parameterize the density function: Gamma(*θ*|*a, b*) ≡ *b*^*a*^/Γ(*a* + 1)*θ*^*a*−1^ exp(−*bθ*). We assume smooth a prior distribution for the *ρ* and *λ* parameters, namely *ρ*_*j*_, *λ*_*gi*_ ∼ Gamma(1, 1). A smaller value for the hyperparameters, such as Gamma(10^−4^, 10^−4^), could encourage the effect of prior distributions vanish; however, we found it often results in numerically-unstable posterior estimation when RNA-seq samples are shallowly sampled.

*Remark:* For the gene parameter *λ*_*gi*_, if we defined its distribution: *λ*_*gi*_ ∼ Gamma(*ϕ*^−1^, *ϕ*^−1^/*μ*_*gi*_), we would have 𝔼[*λ*] = *μ* and 𝕍[*λ*] = *μ*^2^*ϕ*. Integrating out the uncertainty over *λ*, we derive the following negative binomial model:

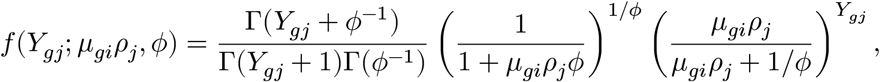

which preserves the characteristic quadratic relationship between the mean and variance: 𝕍[*Y*] = 𝔼[*Y*] + 𝔼[*Y*]^2^ *ϕ*.

#### Variational Bayes for parametric inference

We estimate the posterior distribution of *λ*_*gi*_ and *ρ*_*j*_ by minimizing Kullback-Leibler divergence between the joint likelihood *ℒ* ≡ ∏_*gj*_ *f*(*Y*_*gj*_; *λ*_*gi*_, *ρ*_*j*_)*f*(*λ*)*f* (*ρ*) and the fully-factored variational distributions,^48^ *q*(*λ*) = Gamma(*λ*|*α*_*λ*_, *β*_*λ*_) and *q*(*ρ*) = Gamma(*ρ*|*α*_*ρ*_, *β*_*ρ*_). We can quickly reach convergence by alternating the following update equations:

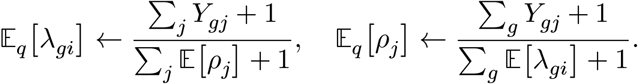

Here, we first initialize 𝔼[*ρ*_*j*_] = 1 for all *j*, and add pseudo-count 1 on both numerators and denominators because of the prior distribution of *ρ* and *λ*.

### Counterfactual confounder adjustment for differential expression analysis

#### Step 1 (Imputation of potential outcomes by Poisson regression)

We assume binary treatment assignment and denote disease assignment (or nature’s treatment) by *W* ∈ {0, 1}. We denote an individual have suffered from a disease by *W* = 1 and the healthy one by *W* = 0. For clarity, we introduce the potential outcome notations to the gene expression variables. Let 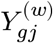 be gene expression of a gene *g* in a cell *j* if this expression value was observed from an individual with a disease label *W* = *w*. For a disease individual, *Y* ^(1)^ is the same as observed *Y* value, but *Y* ^(0)^ is unknown, requiring counterfactual inference; for the opposite case, a healthy individual, *Y* ^(0)^ is observed, but *Y* ^(1)^ is counterfactual. To proceed, we assume the following causal assumptions:^18,20,49^ (1) The disease assignment mechanism (*W*) is unconfounded with potential outcomes *Y* ^(0)^, *Y* ^(1)^, conditioning on some covariates *X*. (2) There is sufficient overlap between the case and control cells with respect to the covariates *X*. In other words, in almost every *X* = *x*, we have 0 < *P* (*W* = 1|*X* = *x*) < 1.

How do we find the counterfactual *Y* ^(1−*w*)^ for the observed *Y* ^(*w*)^? We construct feature vectors for potential outcome prediction by searching *k*-nearest neighbours (NN) from the cells belonging to the opposite conditions. To avoid the curse of dimensionality, we first perform spectral decomposition of single-cell data matrix and efficiently search *k*-NN on the spectral domain with hierarchical hashing algorithm.^50^ Using these counterfactually-matched cells, we construct feature matrix with each element 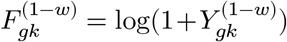 and quickly estimate regression coefficients *β*’s in the Poisson regression by coordinate-wise descent method:^51^

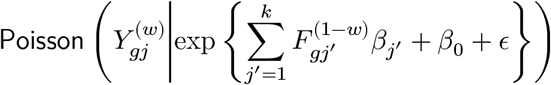

where *β*_0_ captures the intercept term.

Given the optimized coefficients, we predict the potential outcome 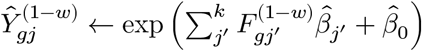, ignoring the residual errors (*ϵ*). We also considered a non-parametric imputation method which takes weighted average over the matched cells.^30,52,53^ Although such non-parametric methods are frequently used in single-cell data analysis, we found that Poisson regression yields more robust performance with fewer neighbouring cells than the other kNN-based imputation methods.

#### Step 2 (Identification of potential confounding effects)

After the matching followed by the regression, we have observed 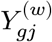 and counterfactual 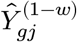. By construction, one of them carry disease-relevant effects unlike the other one. However, both of them can provide disease-invariant information that implicate potential confounding effects, denoted by *μ*_*gi*_ for a gene *g* and individual *i*:

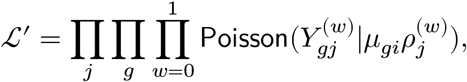

where we introduce the conditional-specific sequencing depth parameters *ρ*^(*w*)^. However, note that *μ*_*gi*_ is shared and label-invariant.

We estimate the posterior mean of *μ*_*gi*_ by variational Bayes by alternating the following update equations until convergence:

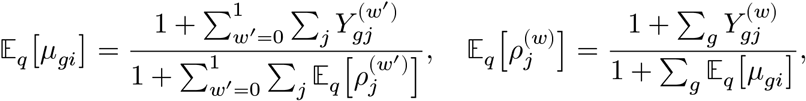

for all *w* ∈ {0, 1}.

#### Step 3 (Confounder adjustment)

While fixing the value 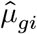 to its (variational) posterior mean 𝔼_*q*_[*μ*_*gi*_], we redeem the confounder-adjusted mean parameters *δ*_*gi*_, by maximizing the data likelihood:

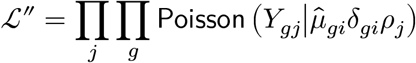

Again, the posterior distributions are found by alternating the following update equations:

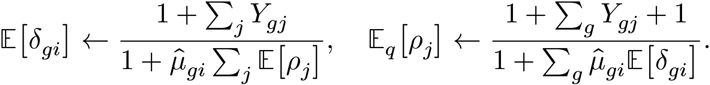

Since the *δ*_*gi*_ variable follows Gamma distribution, we also have

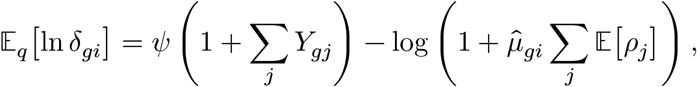

where *ψ*(·) is the digamma function, and approximate its variance,

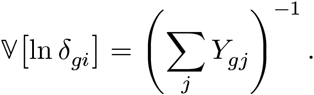

See the supplementary method for the derivation of the Gaussian approximation of Gamma distribution.

### Cell type annotation by constrained mixture of von Mises-Fisher

We classify a cell type of 70,634 cells based on the prior knowledge of cell-type-specific 2,648 marker genes on 8 brain cell types.^29^ Using 1,726 genes present in our data, we construct a normalized vector **m**_*j*_ for each cell with the dimensionality *d* = 1726 and ‖**m**_*j*_‖ = 1. Additionally we define a label matrix *L* to designate the activities of the marker genes to the relevant cell types. Each element *L*_*gk*_ takes 1 if and only if a gene *g* ∈ [*d*] is active on a *k* ∈ [8] cell type, otherwise we set *L*_*jk*_ = 0. We assume that each normalized vector **m**_*j*_ follows von Mises-Fisher (vMF) distribution with cell type *k*-specific mean vector *θ*_*k*_ with the concentration parameter *κ*, shared across all the cell types:

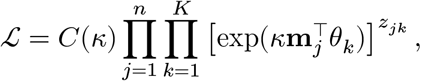

where *n* = 70634 for the cells and *K* = 8 for the cell types. Here, we introduce *z*_*jk*_, an indicator variable to mark the assignment of a cell *j* to a cell type *k*. Our goal is to estimate the posterior probability of *z*_*jk*_ = 1 by stochastic expectation maximization (EM) algorithm. In the E-step, we simply sample the latent membership *z*_*jk*_ from the discrete distribution proportional to 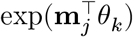. In the M-step, we maximize the mean and concentration parameters with the cell type constraints *L*:

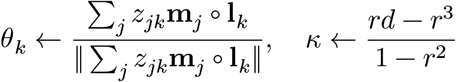

where *r* = ‖ ∑_*j*_ *X*_*j*_‖/*N*. Derivation for the optimization of *κ* can be found in the previous work on von Mises Fisher mixture model54.

## Data Availability

Contact Yongjin Park for the questions on analysis Scripts (https://github.com/YPARK/cocoa_paper). C++ programs are publicly available at GitHub (http://github.com/ypark/mmutil).

http://github.com/ypark/mmutil

## Acknowledgement

We thank Liang He, Matthew Lincoln, and Tomokazu Sumida for biological inspiration, which later led to adopting von Mises-Fisher model for cell type annotations. We also thank Abhishek Sarkar for helpful discussion on individual-level quantification model for single-cell RNA-seq data. We owe a debt of gratitude to anonymous reviewers for constructive feedback that significantly improved the exposition of this manuscript.

The authors declare no competing interests.

## Supplementary methods

### Local Gaussian approximation of Gamma distribution

We approximate the distribution of ln *λ* by constructing a local quadratic approximation of the original log-probability density function:

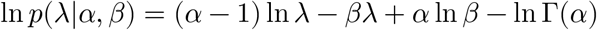

Letting *ϕ* = ln *λ*, we can rewrite the above as:

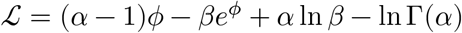

At some 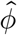, we can find a quadratic form:

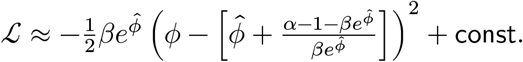

Setting 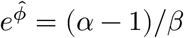 (the mode of Gamma distribution), we have

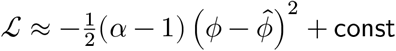

Finally, we have

*p*(*ϕ*|*α, β*) ≈ *𝒩*(*ϕ*∣ln((*α* − 1)/*β*), (*α* − 1)^−1^). In our case, we assumed *λ* ∼ Gamma(1, 1) *a priori* and only derived approximate Gaussian whenever we have at least 1 read per individual, therefore *α* > 1. However, if 0 < *α* ≤ 1, we can approximate the Gaussian at *λ* = *α*/*β*, and this results in *p*(*ϕ*|*α, β*) ≈ *𝒩*(*ϕ*∣ln(*α*/*β*), *α*^−1^).

### Derivation of average disease effect across individuals (meta-analysis)

From the above, we derived the posterior distribution of *ϕ*_*i*_(≡ ln *λ*_*i*_) variables. Let *η*_*i*_ = ln((*α*_*i*_ − 1)/*β*) and 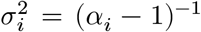. Then we have 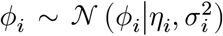. We can find another variational distribution 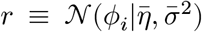 averaging over all these individual-level posterior distributions by optimizing the following Kullback-Leibler divergence:

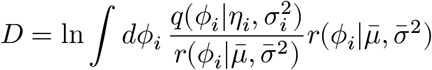

By Jensen’s inequality,

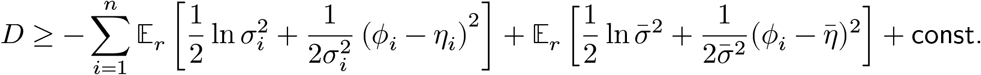

Optimizing this with respect to 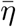 and 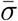, we have:

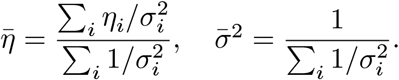

## Figures

**Supplementary Figure 1.**
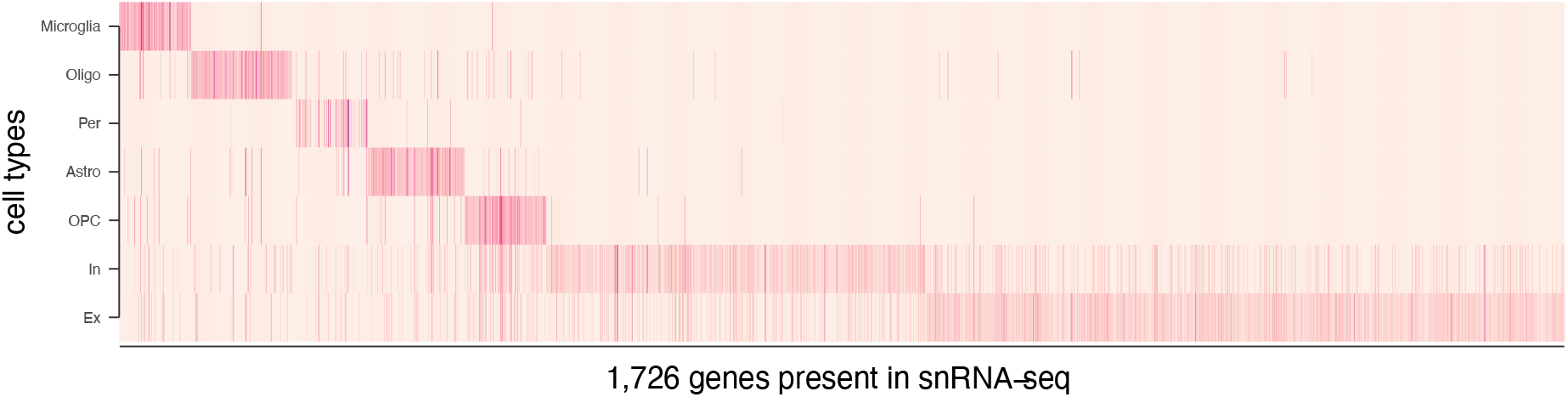
Average cell-type-specific profiles of 1,726 marker genes.

**Supplementary Figure 2.**
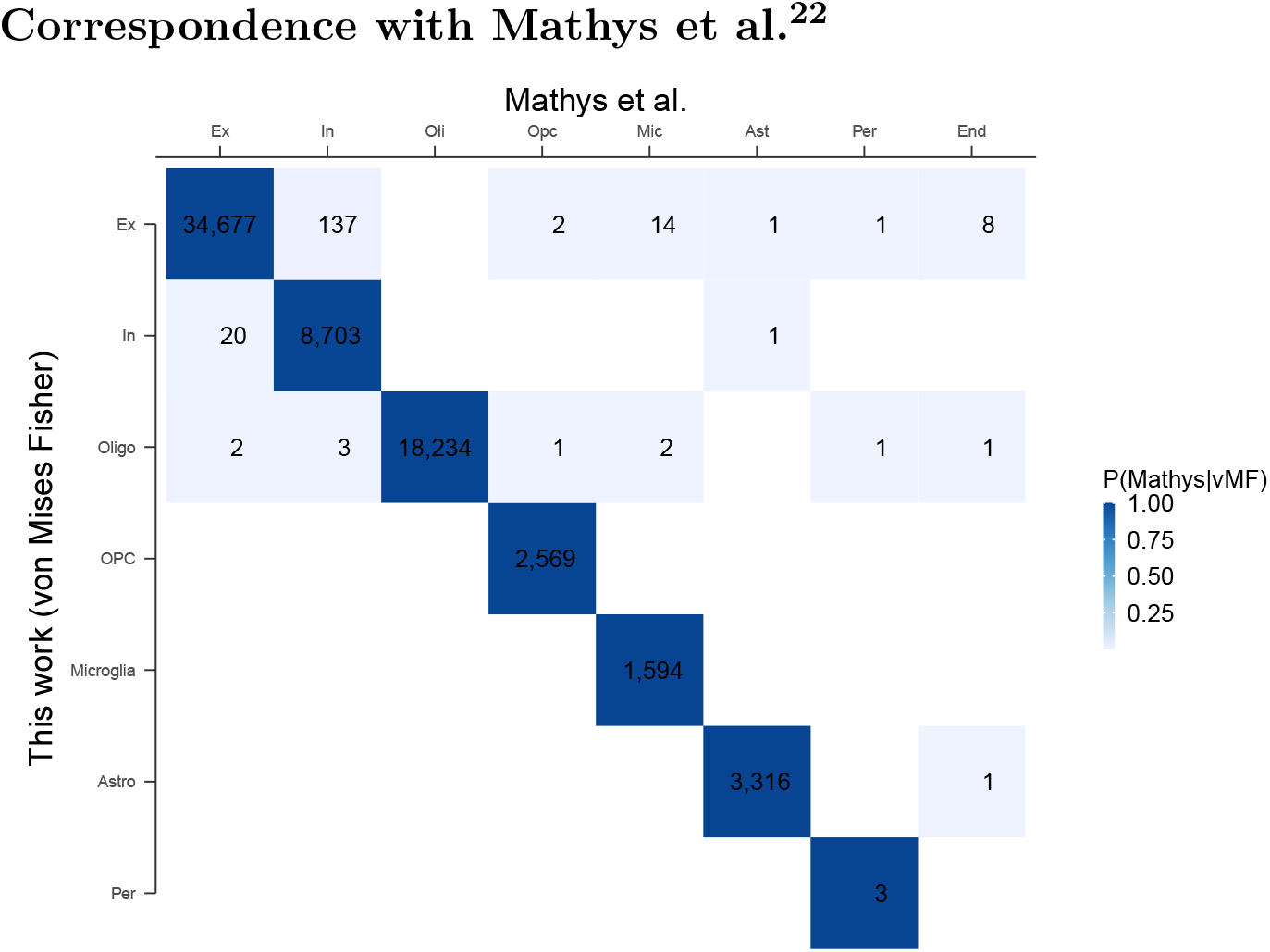
Correspondence with Mathys et al.^22^.

**Supplementary Figure 3.**
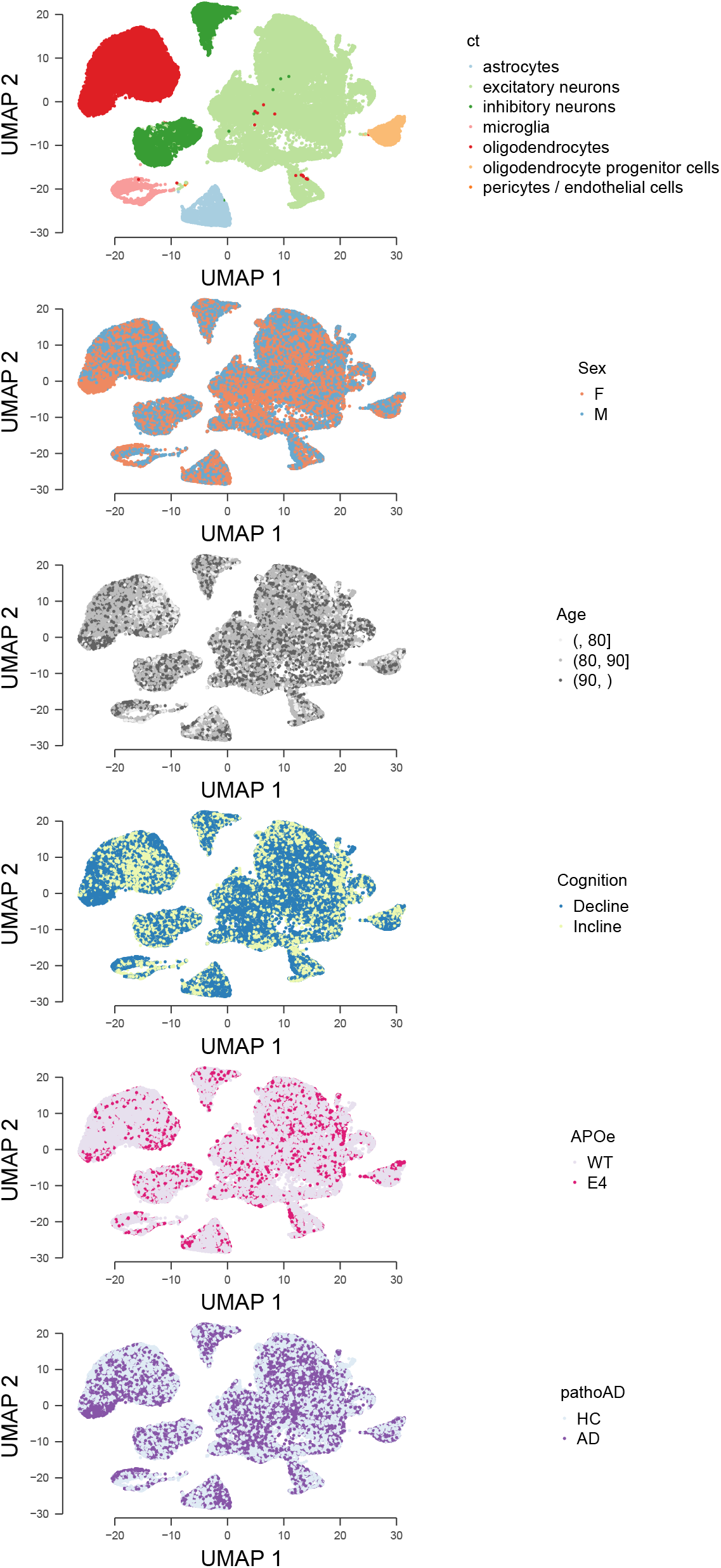
The annotations of the major neuronal and glial cell types are not biased by known biological variables.

**Supplementary Figure 4.**
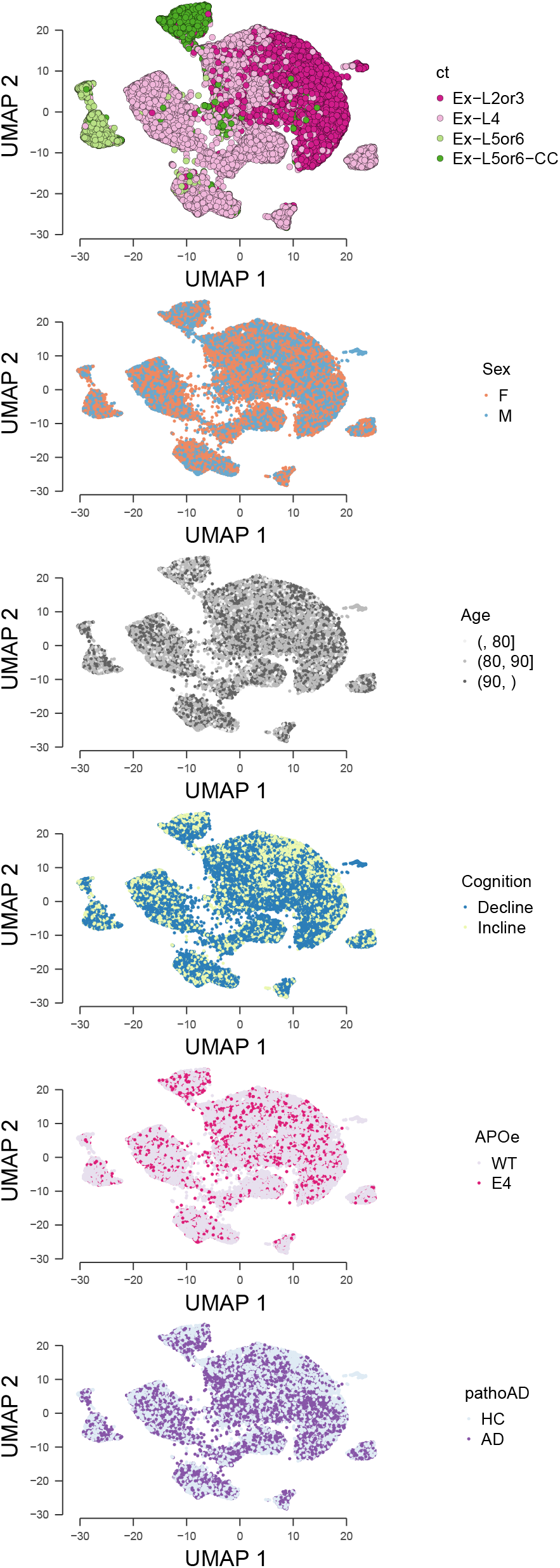
The annotations of the excitatory neuron types are not biased by known biological variables.

**Supplementary Figure 5.**
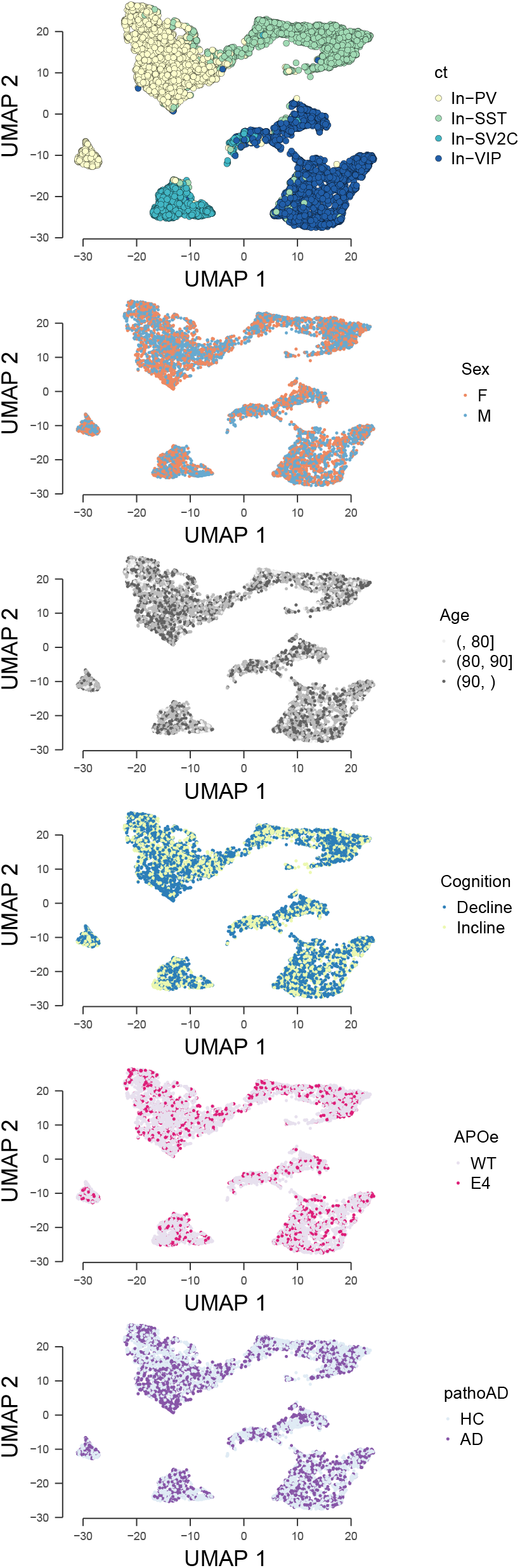
The annotations of the inhibitory neuron types are not biased by known biological variables.

**Supplementary Figure 6.**
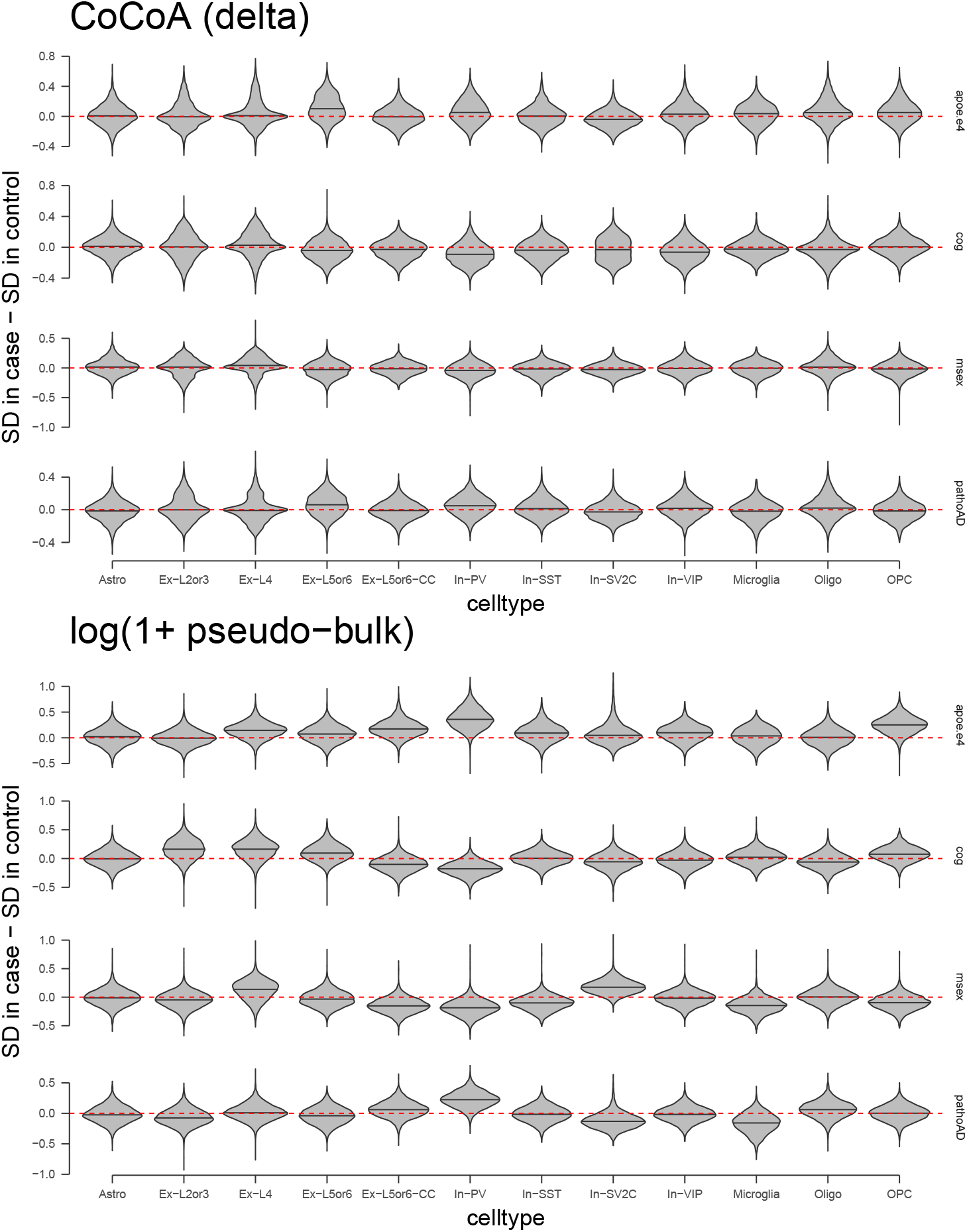
CoCoA algorithm does not create a skewed distribution of variance.

**Supplementary Figure 7.**
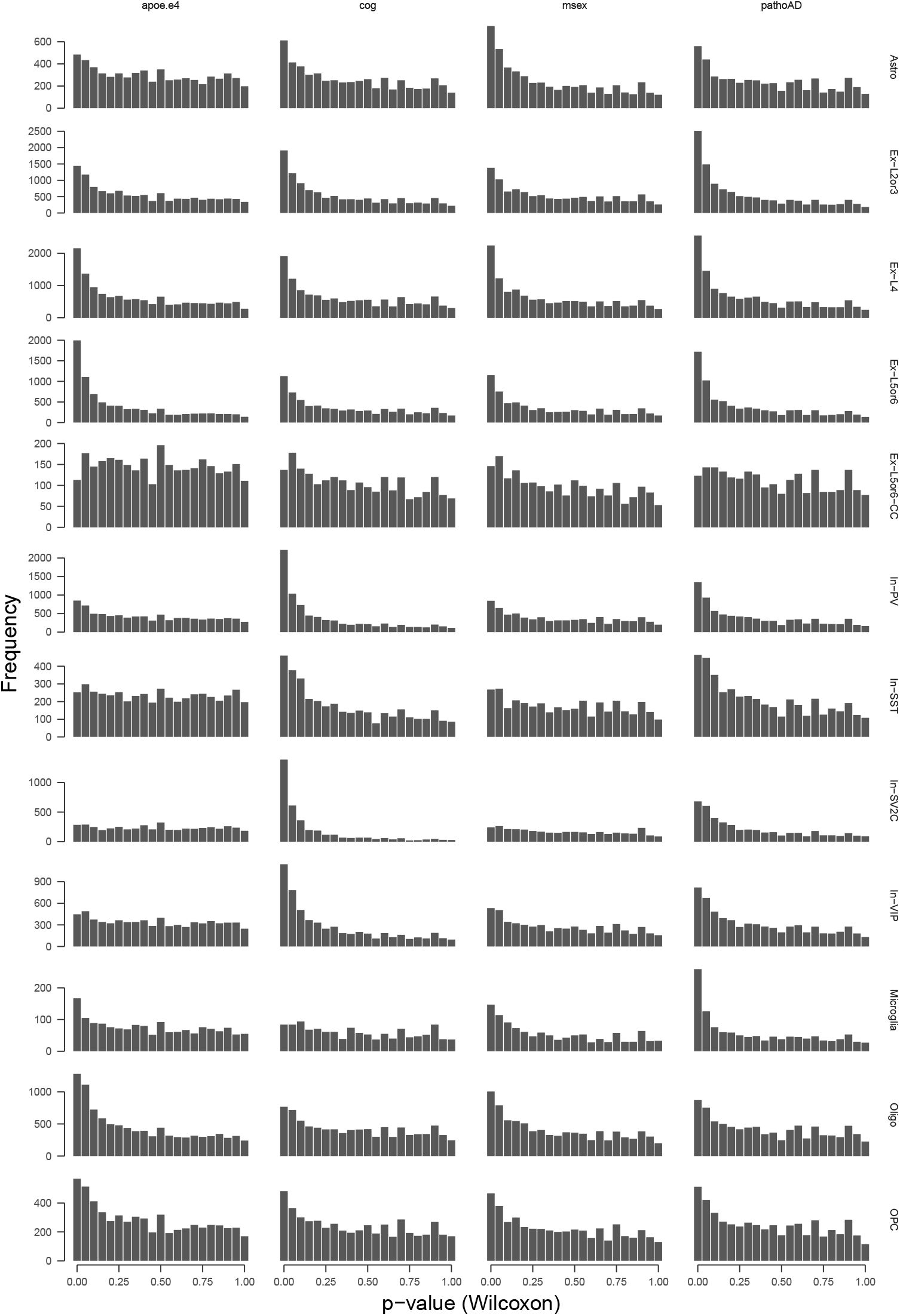
Histogram of p-value distributions.

**Supplementary Figure 8.**
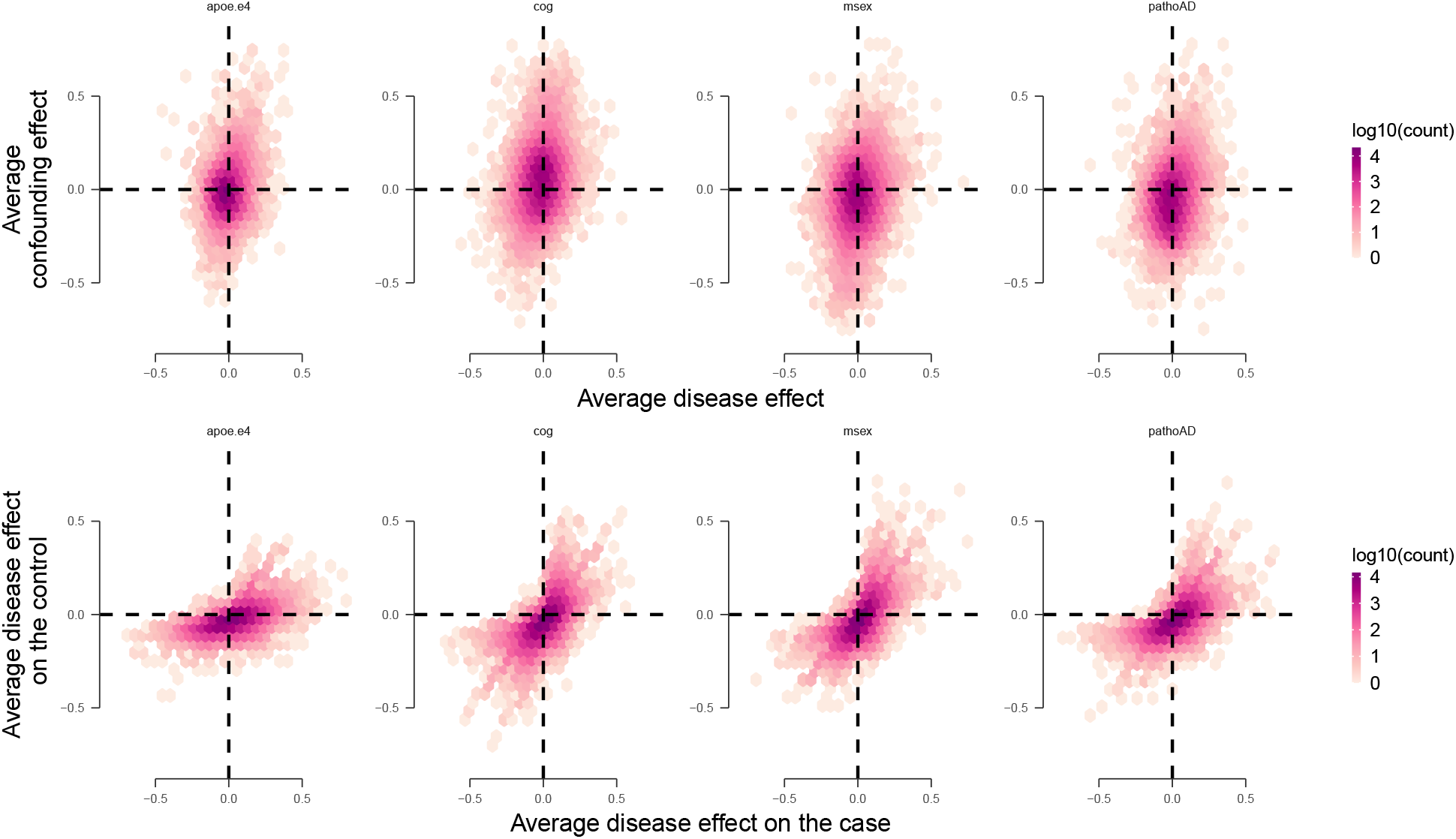
**Top**: Correlations between the average disease effects (ADE) and gene-level associations with the confounding factors. **Bottom**: Correlations between the average disease effects computed on the disease cohort (ADD) and the average disease effects computed on the control cohort (ADC).

## Notes

### Competing Interest Statement

The authors have declared no competing interest.

### Funding Statement

This work was supported by NIH grants U01NS110453, U24-HG009446, and U01-RFA-HG009088 (MK). We also acknowledge generous supports from the BC Cancer Foundation, Project ID 1NSRG048 (YPP).

### Author Declarations

The results published here are in whole or in part based on data obtained from the AD Knowledge Portal (https://adknowledgeportal.synapse.org). Study data were provided by the Rush Alzheimer's Disease Center, Rush University Medical Center, Chicago. Data collection was supported through funding by NIA grants RF1AG57473 (single nucleus RNAseq) and the Illinois Department of Public Health (ROSMAP). The Religious Orders Study and Rush Memory and Aging Project were approved by an IRB of Rush University Medical Center.

